# Streamlining Eligibility Assessment for Alzheimer’s Disease-Modifying Therapies: Prediction of MMSE Scores Using the Digital Clock and Recall

**DOI:** 10.64898/2026.03.03.26347542

**Authors:** Ali Jannati, Claudio Toro-Serey, Marissa Ciesla, Emma Chen, John Showalter, David Bates, Alvaro Pascual-Leone, Sean Tobyne

## Abstract

**Introduction:** The eligibility of anti-amyloid disease-modifying therapies (DMTs) and their integration into clinical practice in some institutions requires a specific range of Mini-Mental State Examination (MMSE) scores. Reliance on this pencil-and-paper psychometric instrument imposes operational burdens and risks perpetuating health disparities due to the test’s known educational and cultural biases. This study evaluates the efficacy of the Digital Clock and Recall (DCR™) – a rapid, FDA-listed digital cognitive assessment – to crosswalk to MMSE scores using machine learning, thereby offering a faster, scalable, and equitable mechanism for patient triage.

**Methods:** We conducted a retrospective analysis using data from the multi-site Bio-Hermes-001 study (NCT04733989, N=945). Participants were clinically classified as cognitively unimpaired, mild cognitive Impairment, or probable Alzheimer’s dementia. We trained a Poisson elastic net regression model using age and multimodal digital features derived from the DCR (including drawing kinematics and voice acoustics) to predict MMSE scores. The model was tested for generalizability using an independent external validation cohort from the Apheleia study (NCT05364307, N=238).

**Results:** The machine learning model predicted MMSE scores with a root mean squared error (RMSE) of 2.31 in the training cohort. This error margin falls within the established test-retest reliability range of the manual MMSE itself (2–4 points), suggesting the prediction is statistically non-inferior to human administration. External validation in the Apheleia cohort demonstrated robust generalizability (RMSE = 2.62). Crucially, the model exhibited demographic fairness, maintaining consistent accuracy across Race (White RMSE = 2.34; Non-White RMSE = 2.14) and Ethnicity (Hispanic RMSE = 2.26; Non-Hispanic RMSE = 2.31).

**Discussion:** Machine learning can leverage multimodal features from the DCR to accurately and equitably crosswalk to MMSE scores in support of current guidelines, transforming a time-intensive manual test into a rapid, automated assessment. By deploying this “digital triage” engine, where traditional assessments are still used for DMT eligibility, healthcare systems can streamline the identification of DMT-eligible patients, reduce specialist referral bottlenecks, and ensure that access to life-altering therapies is determined by pathology rather than demography.

## Introduction

The recent regulatory approval of anti-amyloid disease-modifying therapies (DMTs), specifically lecanemab (Leqembi®) and donanemab (Kisunla®), marks a historic inflection point in the management of patients with Alzheimer’s disease (AD) (1,2). For the first time, clinicians possess pharmacological treatments capable of slowing the clinical progression of mild cognitive impairment (MCI) or early dementia due to AD by targeting underlying amyloid pathology. However, the determination of eligibility for DMTs and their successful integration into clinical practice at some institutions is currently impeded by the reliance on legacy cognitive-screening tests, most notably the Mini-Mental State Examination (MMSE). The pivotal clinical trials that ultimately supported the Food and Drug Administration (FDA) approval of lecanemab and donanemab, i.e., CLARITY AD and TRAILBLAZER-ALZ 2, utilized specific MMSE score ranges (23–29 and 20–28, respectively) as key inclusion criteria, positioning these scores as indicators of eligibility post-market (3,4). Consequently, millions of older adults must be screened to identify the subset eligible for confirmatory biomarker testing (e.g., amyloid PET or CSF analysis) and subsequent therapy. The current primary care infrastructure is ill-equipped to handle this volume. The MMSE, while commonly used, requires 10–15 minutes of direct clinician administration – a prohibitive cost in primary care settings where visit times average less than 15 minutes (5). Furthermore, the test suffers from significant inter-rater variability and “ceiling effects” that mask early impairment in high-functioning individuals (6,7).

More critically, the MMSE is structurally biased against individuals from underrepresented racial and ethnic groups and those with lower educational attainment. Studies have consistently demonstrated that Black and Hispanic older adults are more likely to be misclassified as impaired by the MMSE compared to White counterparts with similar levels of pathology, largely due to cultural and educational nuances in test items (8,9). Relying on such a biased instrument for gatekeeping high-value care threatens to exacerbate systemic healthcare inequities, potentially denying life-altering treatment to disadvantaged populations.

To address these challenges, the field is increasingly turning to digital cognitive Assessments (DCAs). Linus Health’s Digital Clock and Recall (DCR™) represents a technological evolution of the Mini-Cog (10), utilizing a digitizing, commercially available tablet to capture the process of completing a cognitive task with millisecond precision, as opposed to only considering the final product. By analyzing nearly 2000 process-based kinematic, temporal, acoustic, and speech features, such as processing speed, hesitation, and motor planning, the DCR offers a more sensitive, objective, and less biased measure of cognitive function than traditional pen-and-paper scores (10–15). However, the clinical utility of such DCAs depends on their ability to interface with existing regulatory frameworks and the way clinicians adopt them in practice. Despite an ever-growing body of literature detailing the superiority of DCAs over traditional assessments, clinicians may be reluctant to rapidly replace long-standing, familiar instruments. Providing a validated score “crosswalk” between a new DCA and legacy measures (e.g., MMSE or MoCA) can improve interpretability and support incremental adoption while clinical workflows evolve to leverage newer, superior tools (10,11,16–18).

This study presents the development and validation of a machine learning (ML) model capable of predicting MMSE scores using multimodal features derived from the DCR. Leveraging data from a large multi-site study and an external validation cohort, we demonstrate that an ML model based on DCR features can predict MMSE scores with accuracy comparable to the test-retest reliability of the MMSE itself. By providing a rapid, automated, scalable, and equitable crosswalk for the MMSE, this innovation is designed to streamline “digital triage” for clinicians who currently rely on MMSE scores, improving referral decisions, optimizing access to specialist care, and supporting broader, fairer access in the new era of Alzheimer’s therapeutics.

## Methods

Data were obtained from 945 participants enrolled in the multi-site Bio-Hermes-001 study (BH; ClinicalTrials.gov ID: NCT04733989). The study was coordinated by the Global Alzheimer’s Platform Foundation (GAP), with protocols implemented at community-based clinical trial sites. This was an observational study designed to build a biomarker database by collecting blood-based biomarker measures and comparing them to brain amyloid status determined by amyloid PET or CSF. Participants also completed a battery of cognitive assessments, including a mix of traditional assessment and DCAs. Participants or their legally authorized representatives provided written informed consent prior to participation.

For external validation, we used data from 238 participants in the multisite Apheleia-001 study (ClinicalTrials.gov ID: NCT05364307), coordinated by the Global Alzheimer’s Platform Foundation (GAP) in collaboration with AbbVie, Inc. The study evaluated screening strategies, particularly blood-based biomarker testing, intended to reduce screen failures in Alzheimer’s disease therapeutic clinical trials. The objective was to identify and characterize individuals reporting memory concerns and/or cognitive impairment using demographics, clinical history, brief cognitive measures, and blood-based biomarkers, thereby increasing the likelihood of trial randomization (19). Written informed consent was obtained from each participant or their legally authorized representative prior to enrollment.

### Participants and Study Cohort

#### Bio-Hermes-001 study

Full enrollment criteria have been reported previously (13,20). The sample had a mean (±SD) age of 72.0 (±6.7) years; 57% were female; 10% identified as Hispanic or Latino; 85% as White, and 11% as Black or African American. Participants had a mean (±SD) of 15.0 (±2.7) years of education, and English was the primary language for all participants (Table 1). Participants were prospectively categorized by the study team as cognitively unimpaired (CU; n = 402), mild cognitive impairment (MCI; n = 298), or probable Alzheimer’s dementia (pAD; n = 242). Group assignments were determined at the screening visit (Visit 1) using MMSE performance (21), the Rey Auditory Verbal Learning Test (RAVLT) (22), and the Functional Activities Questionnaire (FAQ) (23), together with a clinical interview and medical record review conducted by study site personnel. Alternatively, participants were assigned to the MCI or pAD cohorts based on a clinical diagnosis made within three months of Visit 1. Detailed classification criteria are provided in prior work (see Supplementary Materials in (13)). Analyses in the present study used only Visit 1 data, including the DCR. Performance on the DCR was not considered during cohort assignment and authors remained blinded to cohort membership until database lock.

**Table 1.** Demographics for Bio-Hermes-001 and Apheleia-001 studies. The BH study sample was used to train the model, and the Apheleia sample was used to externally validate it. No cognitive cohorts were available for the Apheleia study.

| Measure / Study | Bio-Hermes-001 |  |  |  | Apheleia-001 |
| --- | --- | --- | --- | --- | --- |
|  | Healthy | MCI | Probable AD dementia | No Diagnosis | – |
| N | 402 | 298 | 242 | 3 | 238 |
| Sex (% Female) | 62 | 54 | 53 | 67 | 65 |
| Age (Median [SD]) | 70 (6.48) | 72 (6.77) | 75 (6.25) | 76 (4.73) | 68 (10.35) |
| Years of Education (Median [SD]) | 16 (2.45) | 16 (2.76) | 15 (2.99) | 18 (2.00) | 15 (2.65) |
| Ethnicity (% Hispanic) | 7 | 11 | 14 | 0 | 19 |
| Race (% White) | 87 | 86 | 84 | 100 | 76 |

#### Apheleia-001 study

Eligible participants were 50 to 90 years old (inclusive) and had progressive cognitive concerns reported by the participant or a caregiver. Individuals with a known or self-reported negative amyloid PET scan within the prior 24 months were excluded. Participants were also excluded if they had experienced a stroke within six months of prescreening or had another neurological disorder (aside from AD) that investigators judged could be contributing to cognitive impairment. The sample had a mean (±SD) age of 68.0 (±10.4) years; 65% were female; and 19% identified as Hispanic or Latino. Racial composition was 76% White, 15% Black or African American, 4% Asian, and 4% Other. Participants had a mean (±SD) of 15.0 (±2.7) years of education, and English was the primary language for all participants. All participants completed both the DCR and the MMSE. Beyond administration of the cognitive measures, the study protocol did not assign an overall cognitive diagnostic classification to participants.

### Digital Clock and Recall (DCR)

Linus Health’s DCR (11–13) is an FDA-listed Class II software as a medical device and a multimodal, ML–enabled evolution of the Mini-Cog (10). The three-minute assessment is completed on an iPad using an Apple Pencil and can be administered by medical or research assistants without specialized training.

The DCR is administered in three consecutive steps:

1. The assessment begins with an *Immediate Recall* task: three unrelated words are read aloud, and the participant repeats them to confirm encoding. Responses are spoken and recorded without a time limit. This step is not included in the overall DCR Score, but helps assess attention/hearing and establishes a baseline for the later delayed recall.
2. Immediately after word encoding, the participant completes the *digital clock drawing test* [DCTclock™(24,25)] on an iPad in two sequential conditions. In the Command Clock condition, they draw a clock from memory, placing the numbers correctly and setting the hands to 11:10 without any template or visual cues. In the Copy Clock condition, they reproduce an on-screen clock set to the same time. Both drawings are completed with a stylus while the software records completion time and captures detailed drawing behavior data in addition to the final images. Together, these tasks probe visuospatial and visuoconstructional ability, planning and executive function, processing speed, and motor control.
3. After the clock drawing tasks, the participant is asked to recall the original three words. Responses are spoken and recorded, while the application logs both response content and captures detailed acoustic and speech features. This *Delayed Recall* component assesses verbal episodic memory, which is often sensitive to early AD pathology, and completes the DCR administration.

### Data acquisition and feature extraction in the DCR

The DCR passively captures high-resolution drawing and speech data to derive a broad set of quantitative “process features.” During clock drawing, the system records time-stamped pen trajectories, enabling the extraction of velocity profiles, acceleration, stroke length, pen speed, total drawing duration, and time on-task vs. time off-tablet. It also derives temporal features, including pauses and latencies such as initiation time, inter-stroke intervals, and stage-specific latencies. From the final drawings, it computes spatial/organizational measures including clock face symmetry and circularity, numeral placement, clock hand placement and proportions, and overall vertical and horizontal positioning on the screen (24,26). In parallel, the DCR records spoken responses during recall to extract response latency, total duration, pauses, and acoustic features such as pitch, jitter, shimmer, and speaking rate (12,13). Together, these modalities support the generation of roughly ∼2000 features used by ML models for detection beyond traditional scoring.

### DCR Scoring

DCR scoring is fully automated. The delayed recall audio is transcribed via automatic speech recognition, and the recognized words are matched to the three target words for a 0–3 score (1 point per exact match). Immediate recall is recorded but not included in the score, serving primarily as an encoding/attention/hearing check, and can be reviewed clinically.

The clock drawing portion is scored automatically using a combined machine learning and rules-based engine. The system first “parses” the drawing by classifying each pen stroke into clock components (e.g., clock face, numerals, hour/minute hands, and extraneous marks/self-corrections) using both spatial layout and the temporal sequence of strokes. This stroke-level labeling, trained on large sets of annotated clocks and reinforced with hierarchical rules, supports robust scoring even for messy drawings and enables the extraction of cognitively informative timing and process metrics. Development, validation, and biomarker associations for the DCTclock are detailed in several prior studies (24–33).

The DCR generates an ML-derived model for accurate identification of cognitive impairment, computed from multimodal features captured during the DCR (13). The result is reported in three tiers: Green indicates no significant impairment detected, Yellow suggests borderline findings warranting monitoring or intervention, and Red indicates a high likelihood of impairment consistent with MCI or dementia. The automated report presents the score and tier, along with playback and component-level breakdown to support rapid clinical interpretation, and has been evaluated in multiple real-world implementation studies (34–41).

### Analytical approach

We split participants into training (70%) and test (30%) sets, ensuring that the distribution of MMSE scores in each subset was similar to the full set (median=27, SD=2.81, kurtosis=-0.19). DCR features were automatically processed after each administration and included 307 drawing and speech DCR metrics (12,13) plus age. Missing data (< 2%) were imputed using the respective median values from the training set. A 10-fold cross-validated weighted Poisson elastic net regression model using glmnet [glmnetUtils package (42)] was trained using the training set, optimizing for alpha and lambda regularization parameters simultaneously per fold. All DCR features and age were given to the model as potential predictors. Given the sparse representation of low MMSE scores in BH, we assigned additional weighting in the model to scores below 23, with a second-highest weight given to scores above 27. The elastic net model, therefore, took the following form, where the Poisson regression was

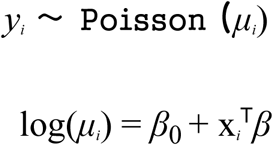

with the weight applied as

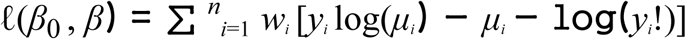

where:

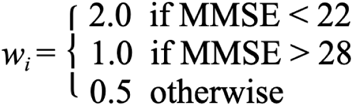

The weights were chosen as the inverse of the rates of scores for each score range. The final form of the penalized model to fit was therefore:

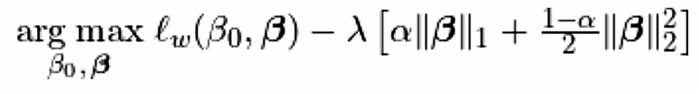

The resulting best model was validated on the test set using the root mean square error (RMSE) and mean absolute error (MAE) on the predicted versus observed MMSE total scores, both overall and stratified by demographics.

Given the smaller sample size of the Apheleia-001 study, we decided not to evaluate performance by demographics, as was done in the larger BH study.

## Results

The final optimized elastic net parameters were 2.98 for alpha and 0.21 for the L1 regularization ratio. The final 15 features selected by the model were related to delayed recall accuracy, total words produced during recall, speech production rates during immediate and delay recall, clock-drawing features related to latency and component placement, and age.

In the BH data set, the DCR model was able to predict MMSE with an RMSE of 2.31 (Figure 1A). RMSE was similar across Gender (male = 2.35; female = 2.27), Ethnicity (Hispanic = 2.26; Non-Hispanic = 2.31), and Race (White = 2.34; Non-White = 2.14) groups (Figure 1A). Based on (6), we found that the short-term reliable change index (RDI) for MMSE (4.10-4.42) was similar to the RDI between our observed and predicted scores (4.22), suggesting that our prediction is within the test-retest reliability range of the MMSE itself within 3 months (43).

**Figure 1.**
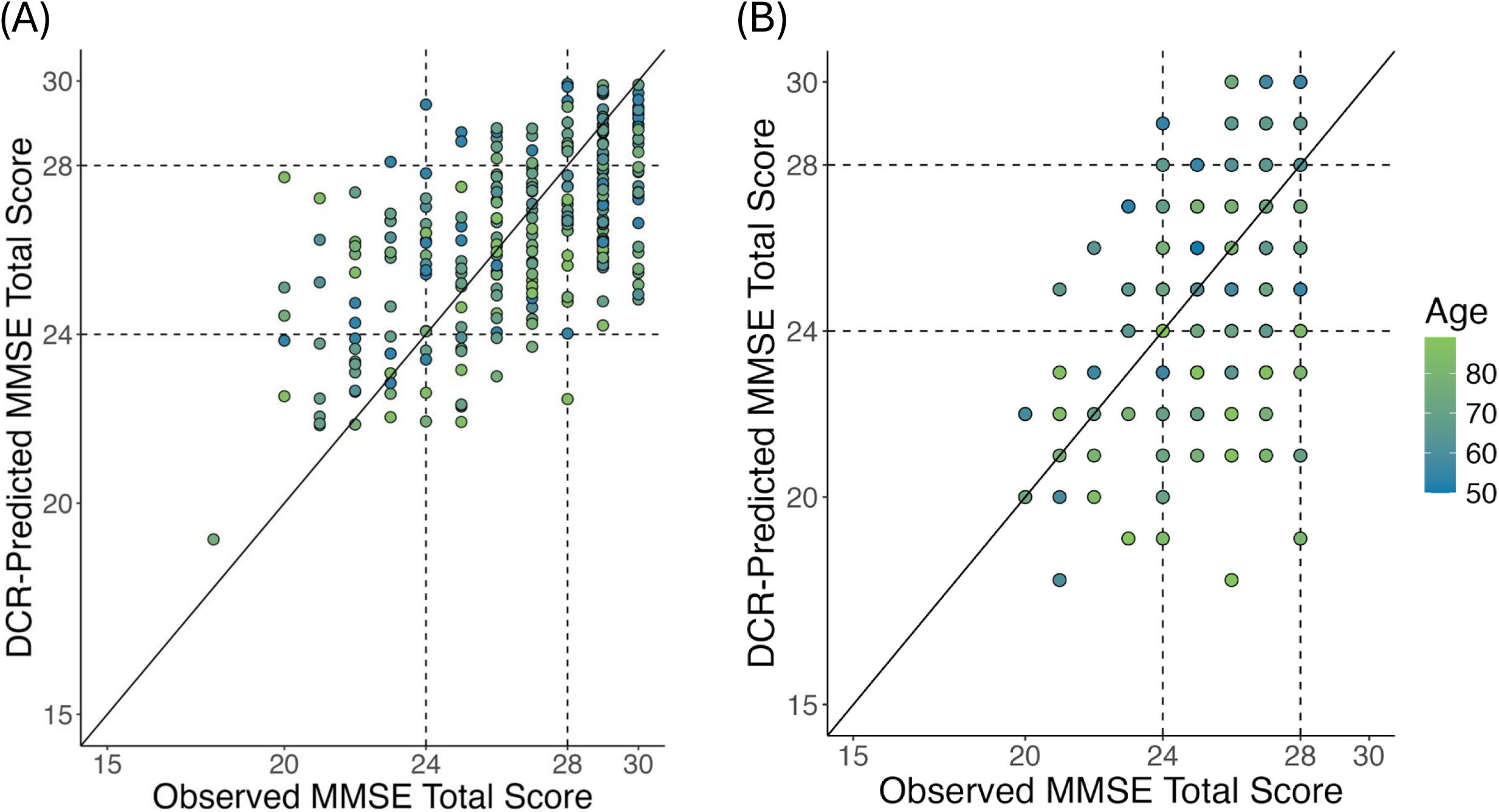
**(A)** Predicted versus actual MMSE total score for an elastic net model of DCR features from the BH data set. Lines indicate MMSE score thresholds denoting putative mild and more severe cognitive impairment. In addition to the strong prediction performance (RMSE = 2.31), the plot shows that model-predicted MMSE scores are not entirely dependent on age. **(B)** Predicted versus actual MMSE total score for an elastic net model of DCR features from the Apheleia cohort. Lines indicate MMSE score thresholds denoting putative mild and more severe cognitive impairment. In addition to the strong prediction performance (RMSE = 2.62), the plot shows that model-predicted MMSE scores are not entirely dependent on age in this dataset either.

We then performed external validation using the Apheleia data set and applying the pre-trained MMSE prediction model without any additional retraining. The previously trained DCR model predicted MMSE scores with an RMSE of 2.62. Figure 1B shows that the relationship between observed and predicted scores was similar to the one in the BH dataset (Figure 1A).

## Discussion

The principal finding of this study is that a machine learning model, leveraging multimodal process-based features from the Digital Clock and Recall (DCR), can predict MMSE scores with a high degree of accuracy (RMSE = 2.31). Additionally, predictive performance was consistent across key demographic groups, suggesting it was not materially influenced by demographic factors. This predictive capability holds important implications for the clinical implementation of novel anti-amyloid therapies. By accurately crosswalking digital motor-linguistic signals into a widely used cognitive staging measure, this approach offers a scalable solution to the diagnostic bottlenecks often contributing to the delay in patient access to disease-modifying therapies (DMTs).

### Benchmarking Accuracy Against the Reference

To properly interpret the model’s performance, it is necessary to contextualize the Root Mean Squared Error (RMSE) of 2.31. While the MMSE remains a key regulatory reference measure for determining eligibility for DMTs, it is psychometrically imperfect. Historical data on the test-retest reliability of the MMSE indicate that scores for the same individual can fluctuate by 2 to 4 points upon repeated administration due to factors unrelated to clinical progression, such as administrator variability, practice effects, and time of day (6,21). Consequently, our model’s prediction error falls within the inherent margin of error of the MMSE test itself. This suggests that the digital prediction is statistically non-inferior to a human re-administration of the MMSE, effectively functioning as an automated “second rater” that eliminates the variability of human scoring (44).

### Addressing the Implementation Crisis for Disease-Modifying Therapies

The FDA approvals of lecanemab and donanemab have created an urgent need for efficient patient identification. Current Appropriate Use Recommendations and payer coverage policies strictly limit these therapies to patients with MCI or early dementia, typically operationalized as an MMSE score of 22–30. This requirement has transformed the MMSE from a general screening tool into one of the main critical gatekeepers for access (45,46).

However, the requirement for manual psychometric testing creates a severe bottleneck. Health system modeling by the RAND Corporation projects that wait times for a specialist evaluation could extend to 50 months due to the shortage of neurologists and the surge in demand (47,48). Over that interval, an individual with Alzheimer’s disease – whose MMSE typically declines by roughly two to four points per year (49) – could lose approximately 8–16 points, progressing from mild to moderate dementia (MMSE < 22) and effectively “aging out” of eligibility before treatment can begin.

The DCR offers a “digital triage” mechanism to mitigate this crisis. Because the test can be self-administered in under 3 minutes by support staff in primary care – requiring no specialized training – it allows for high-throughput screening. By generating a predicted MMSE score immediately, the system can stratify patients who are likely in the “Goldilocks window” for treatment, e.g., recommended MMSE 22–30 for lecanemab (3) and MMSE 20–30 for donanemab (50), thereby prioritizing referrals for confirmatory biomarker testing (e.g., amyloid PET or CSF). This targeted approach optimizes specialist capacity, ensuring that scarce neurological resources are focused on patients most likely to qualify for intervention.

### Mitigating Systemic Bias and Promoting Health Equity

A significant contribution of this study is the demonstration of demographic fairness. The traditional MMSE is known to exhibit bias against individuals from underrepresented racial groups and those with lower educational attainment, often yielding false positives for impairment due to cultural or educational items rather than neuropathology (7–9,51). Such biases in a gatekeeping instrument pose a risk of systemic exclusion, potentially denying life-altering therapies to non-White populations.

Our analysis of the BH sample reveals that the DCR-based prediction model maintains consistent accuracy across racial (White vs. Non-White) and ethnic (Hispanic vs. Non-Hispanic) subgroups. This stability likely stems from the DCR’s reliance on *process-based* features – such as drawing/thinking speed, latency, and motor kinematics – which are more biologically grounded and less culturally mediated than the *content-based* knowledge questions of pen-and-paper tests (11,14,15). By filtering out sociodemographic noise, the digital model provides a more equitable estimate of global cognition, supporting the fair allocation of emerging therapies.

### Advantages Over Traditional Testing

Beyond accurately estimating the MMSE score, the DCR-based approach offers distinct clinical advantages.

● **Sensitivity to Subtle Change:** A prior analysis of the BH dataset found that the DCR is more sensitive than the MMSE for detecting cognitive impairment, identifying subtle impairment even among individuals who score at or near the ceiling on the MMSE (11).
● **Operational Efficiency:** The integration of automated scoring into the electronic health record (EHR) removes the administrative burden of manual scoring and documentation, addressing a primary barrier to cognitive assessment reported by primary care physicians (52). It also mitigates the effects of subjective administration and scoring.
● **Longitudinal Monitoring:** The continuous, granular nature of digital metrics captured by the DCR allows for more precise monitoring of disease trajectory, treatment response, or adverse events than coarse integer-scale tests (31,45).

### Limitations and Future Directions

This study has some limitations. The analysis is cross-sectional; longitudinal validation is required to confirm that changes in predicted MMSE scores track accurately with disease progression and treatment effects over time. Additionally, while the Apheleia cohort provided external validation, further testing in broader community-based primary care populations is necessary to ensure generalizability outside of research-interested volunteers.

## Conclusion

Machine learning applied to multimodal, process-based features of DCR performance can accurately and equitably predict MMSE scores, providing a scalable proxy for traditional testing. As healthcare systems adapt to the era of disease-modifying Alzheimer’s treatments, this tool offers a critical mechanism to streamline eligibility assessment, reduce specialist bottlenecks, and ensure that access to therapy is determined by cognitive impairment and pathology rather than demography.

## Acknowledgements

We thank the participants, organizers, and staff of the Bio-Hermes-001 and Apheleia-001 studies.

## Ethics Statement

The Bio-Hermes-001 and Apheleia-001 studies were performed in accordance with the Declaration of Helsinki and its later amendments. The study procedures were explained to participants verbally and through written informed consent that was approved by the local IRB of each site participating in the GAP consortium for each study. If, in the opinion of the site principal investigator, the participant did not have the capacity to sign the informed consent form, a legally authorized representative was used to grant consent on behalf of the participant.

## Data Availability

The data underlying the findings of this study were collected as part of the Bio-Hermes-001 study (ClinicalTrials.gov ID: NCT04733989) and Apheleia-001 study (ClinicalTrials.gov ID: NCT05364307) and are governed by the Global Alzheimer’s Platform Foundation (GAP) consortium agreement. Data will be made available via the Alzheimer’s Disease Data Initiative (ADDI) Workbench in the future and at the discretion of GAP. All requests for data access should be made directly to GAP. The code used to calculate the reported results is available from Linus Health, Inc. upon reasonable request and with the permission of Linus Health, Inc. Usage restrictions apply to the availability of this code, which is not immediately publicly available.

## Author Contributions

A.J. drafted the introduction and discussion sections and contributed to the methods section. C.T.-S. contributed to the methods section, analyzed the data, and drafted the results section. M.C., E.C., A.P.-L., and S.T. contributed to the introduction and discussion sections. All authors reviewed the manuscript.

